# Bayesian Inference of COVID-19 Spreading Rates in South Africa

**DOI:** 10.1101/2020.04.28.20083873

**Authors:** Rendani Mbuvha, Tshilidzi Marwala

**Affiliations:** School of Statistics and Actuarial Science, University of Witwatersrand, Braamfontien, Gauteng, South Africa; Vice-Chancellor and Principal, University of Johannesburg, Aucklandpark, South Africa

## Abstract

The Severe acute respiratory syndrome coronavirus 2 (SARS-CoV-2) pandemic has highlighted the need for the development of prompt mitigating responses under conditions of high uncertainty. Fundamental to the design of rapid state reactions is the ability to perform epidemiological model parameter inference for localised trajectory predictions. In this work, we perform Bayesian parameter inference using Markov Chain Monte Carlo (MCMC) methods on the Susceptible-Infected-Recovered (SIR) and Susceptible-Exposed-Infected-Recovered (SEIR) epidemiological models with time-varying spreading rates for South Africa. The results find two change points in the spreading rate of COVID-19 in South Africa as inferred from the confirmed cases. The first change point coincides with state enactment of a travel ban and the resultant containment of imported infections. The second change point coincides with the start of a state-led mass screening and testing programme which has highlighted community-level disease spread that was not well represented in the initial largely traveller based and private laboratory dominated testing data. The results further suggest that due to the likely effect of the national lockdown, community level transmissions are slower than the original imported case driven spread of the disease.

## Introduction

The novel coronavirus first manifested in the city of Wuhan, China in December 2019. The disease has subsequently spread around the world, leading to the World Health Organisation (WHO) declaring it a pandemic on 11 March 2020 [1]. In South Africa, by 26 April 2020, 4546 people had been confirmed to have been infected by the coronavirus with 87 fatalities [2].

Governments around the globe, have embarked on numerous efforts to reduce the number of COVID-19 cases [3, 4, 5]. These measures are centred around quarantine and social distancing strategies that seek to separate the infectious population from the susceptible population [5].

These initiatives aim to strategically reduce the increase in infections to a level where their healthcare systems stand a chance of minimising the number fatalities[5]. Some of the key indicators for policymakers to plan appropriately include projections of how much of the population will be affected, how many will require medical attention and whether current containment measures are effective [5].

As the pandemic develops in a rapid and varied manner in most countries, calibration of epidemiological models based on available data can prove to be difficult [6]. The difference between the “confirmed infected” and the “infected” populations escalates this effect due to the high number of asymptomatic cases [1] and constrained testing protocols.

A fundamental issue when calibrating localised models is inferring parameters of compartmental models such as susceptible-infectious-recovered (SIR) and the susceptible-exposed-infectious-recovered (SEIR) that are widely used in infectious disease projections. In the view of public health policymakers, a critical aspect of projecting infections is the inference of parameters that align with the underlying trajectories in their jurisdictions. The spreading rate is a parameter of particular interest which is subject to changes due to voluntary social distancing measures and government-imposed contact bans.

The uncertainty in utilising these models is compounded by the limited data in the initial phases and the rapidly changing dynamics due to rapid public policy changes.

To address these complexities, we utilise the Bayesian Framework for the inference of epidemiological model parameters in South Africa. The Bayesian framework allows for both incorporation of prior knowledge and principled embedding of uncertainty in parameter estimation.

In this work we combine Bayesian inference with the compartmental SEIR and SIR models to infer time varying spreading rates that allow for quantification of the impact of government interventions in South Africa.

## Methods

### Epidemiological Modelling

Epidemiological Modelling of infectious diseases is dominated by compartmental models which simulate the transition of individuals between various stages of illness [7, 8]. We now introduce the Susceptible-Exposed-Infectious-Recovered (SEIR) and the related Susceptible-Infectious-Recovered (SIR) compartmental models that have been dominant in COVID-19 modelling literature [3, 4, 5, 9].

### The Susceptible-Exposed-Infectious-Recovered Model

The SEIR is an established epidemiological model for the projection of infectious disease. The SEIR models the transition of individuals between four stages of a condition, namely:

- being susceptible to the condition,
- being infected and in incubation
- having the condition and being infectious to others and
- having recovered and built immunity for the disease.

The SEIR can be interpreted as a four-state Markov chain which is illustrated diagrammatically in figure 1. The SEIR relies on solving the system of ordinary differential equations below representing the analytic trajectory of the infectious disease [5].

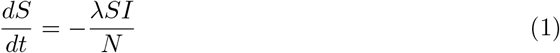

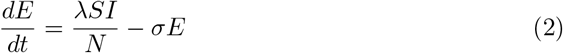

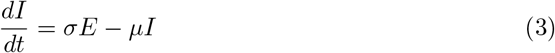

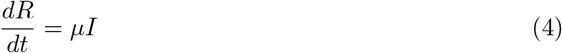

**Fig 1.**
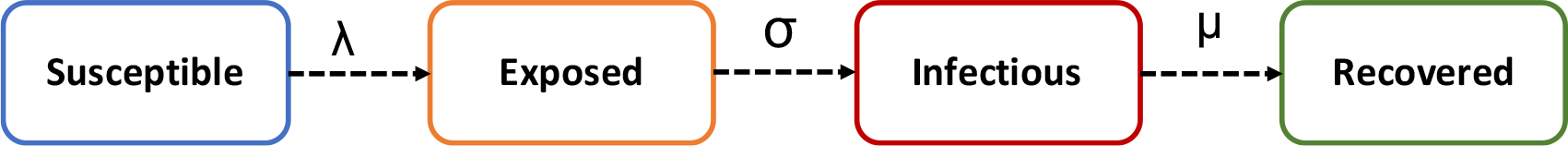
**An Illustration of the underlying states of the Susceptible-Exposed-Infectious-Recovered Model(SEIR)**

Where S is the susceptible population, I is the infected population, R is the recovered population and N is the total population where *N* = *S* + *E* + *I* + *R. λ* is the transmission rate, *σ* is the rate at which individuals in incubation become infectious, and *µ* is the recovery rate. 1*/σ* and 1*/µ* therefore, become the incubation period and contagious period respectively.

We also consider the Susceptible-Infectious-Recovered (SIR) model which is a subclass of the SEIR model that assumes direct transition from the susceptible compartment to the infected (and infectious) compartment. The SIR is represented by three coupled ordinary differential equations rather than the four in the SEIR. Figure 2 depicts the three states of the SIR model.

**Fig 2.**
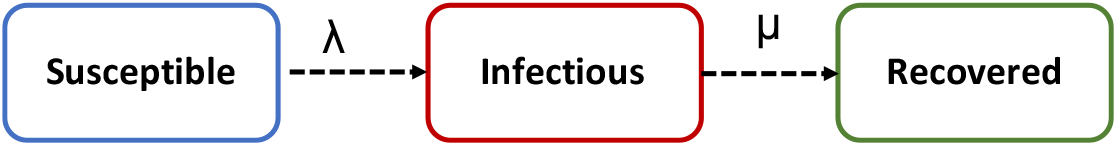
An Illustration of the underlying states of the Susceptible-Infectious-Recovered Model(SIR)

### The Basic Reproductive Number *R*_0_

The contagiousness of a disease is often measured using a metric called the basic reproductive number (*R*_0_). *R*_0_ represents the mean number of additional infections created by one infectious individual in a susceptible population. According to the latest available literature, without accounting for any social distancing policies the *R*_0_ for COVID-19 is between 2 and 3.5[1, 4, 9, 10]. *R*_0_ can be expressed in terms of *λ* and *µ* as:

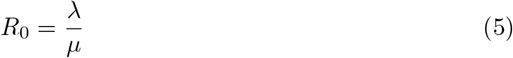

### Extensions to the SEIR and SIR models

We use an extended version of the SEIR and SIR models of [4] that incorporates some of the observed phenomena relating to COVID-19. First we include a delay *D* in becoming infected (*I*^new^) and being reported in the confirmed case statistics, such that the confirmed reported cases CR_*t*_ at some time *t* are in the form [4] :

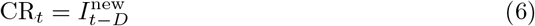

We further assume that the spreading rate *λ* is time-varying rather than constant with change points that are affected by government interventions and voluntary social distancing measures.

### Bayesian Parameter Inference

We follow the framework of [4] to perform Bayesian inference for model parameters on the South African COVID-19 data. The Bayesian framework allows for the posterior inference of parameters which updates prior beliefs based on a data-driven likelihood. The posterior inference is governed by Bayes theorem as follows:

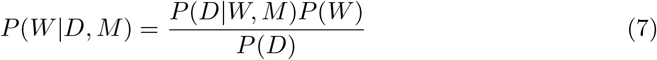

Where *P* (*W*|*D, M*) is the posterior distribution of a vector of model parameters (*W*) given the model(M) and observed data(D), *P* (*D*|*W, M*) is the data likelihood and *P* (*D*) is the evidence.

### The Likelihood

The Likelihood indicates the probability of observing the reported case data given the assumed model. In our study, we adopt the Student-T distribution as the Likelihood as suggested by [4]. Similar to a Gaussian likelihood, the Student-T likelihood allows for parameter updates that minimise discrepancies between the predicted and observed reported cases.

### Priors

Parameter prior distributions encode some prior subject matter knowledge into parameter estimation. In the case of epidemiological model parameters, priors incorporate literature based expected values of parameters such as recovery rate(*µ*), spreading rate(*λ*), change points based on policy interventions etc.

The prior settings for the model parameters are listed in table 1. We follow [4] by selecting LogNormal distributions for *λ* and *σ* such that the initial mean basic reproductive number is 3.2 which is consistent with literature [1, 3, 4, 9, 11]. We set a LogNormal prior for the *σ* such that the mean incubation period is five days. We use the history of government interventions to set priors on change points in the spreading rate. The priors on change-points include 19/03/2020 when a travel ban and school closures were announced, and 28/03/2020 when a national lockdown was enforced. We assume apriori that each intervention reduces the mean spread rates from the Lognormal distributions. Similar to[4] we adopt “weakly-informative” Half-Cauchy priors for the initial conditions for the infected and exposed populations.

**Table 1.** Prior distribution settings for SEIR and SIR model parameters.

| Parameter | Prior Distribution |
| --- | --- |
| Spreading rate $\lambda_0$ | LogNormal(log(0.4),0.5) |
| Spreading rate $\lambda_1$ | LogNormal(log(0.2),0.5) |
| Spreading rate $\lambda_2$ | LogNormal(log(1/8),0.2) |
| Incubation to infectious rate $\sigma$ | LogNormal(log(1/5),0.5) |
| Recovery rate $\mu$ | LogNormal(log(1/8),0.2) |
| Reporting Delay $D$ | LogNormal(log(8),0.2) |
| Initial Infectious $I_0$ | Half-Cauchy(20) |
| Initial Exposed $E_0$ | Half-Cauchy(20) |
| Change Point $t_1$ | Normal(2020/03/18,1) |
| Change Point $t_2$ | Normal(2020/03/28,1) |

### Markov Chain Monte Carlo (MCMC)

Given that the closed-form inference of the posterior distributions on the parameters listed in table 1 is infeasible, we make use of Markov Chain Monte Carlo to sample from the posterior. Monte Carlo methods approximate solutions to complex numerical problems by simulating a random process. MCMC uses a Markov Chain to sample from the posterior distribution, where a Markov Chain is a sequence of random variables *W*_*t*_ such that:

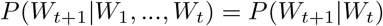

MCMC techniques have been widely used in COVID-19 parameter inference [4, 9]. In this work, we explore inference using Metropolis-Hastings (MH), Slice Sampling and No-U-Turn Sampler (NUTS).

#### Metropolis Hastings (MH)

MH is one of the simplest algorithms for generating a Markov Chain which converges to the correct stationary distribution. The MH generates proposed samples using a proposal distribution. A new parameter state *W*_*t∗*_ is accepted or rejected probabilistically based on the posterior likelihood ratio:

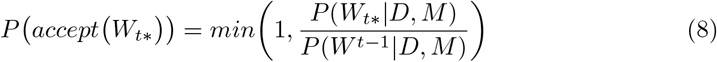

A common proposal distribution is a symmetric random walk obtained by adding Gaussian noise to a previously accepted parameter state. Random walk behaviour of such a proposal typically results in low sample acceptance rates.

#### Slice Sampling

Slice sampling facilitates sampling from the posterior distribution *P* (*W*|*D, M*) by adding an auxiliary variable *u* such that the joint posterior distribution becomes:

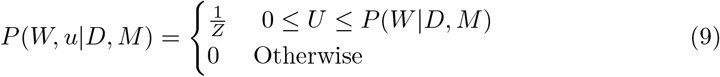

Where *Z* = *P* (*W*|*D, M*)*dW* which is a normalisation constant. Marginal samples for the parameters W can then be obtained by ignoring *u* samples from the joint samples. This process corresponds to sampling above the slice of the posterior density function around a predefined window. Figure 3 shows an illustration of slice sampling.

**Fig 3.**
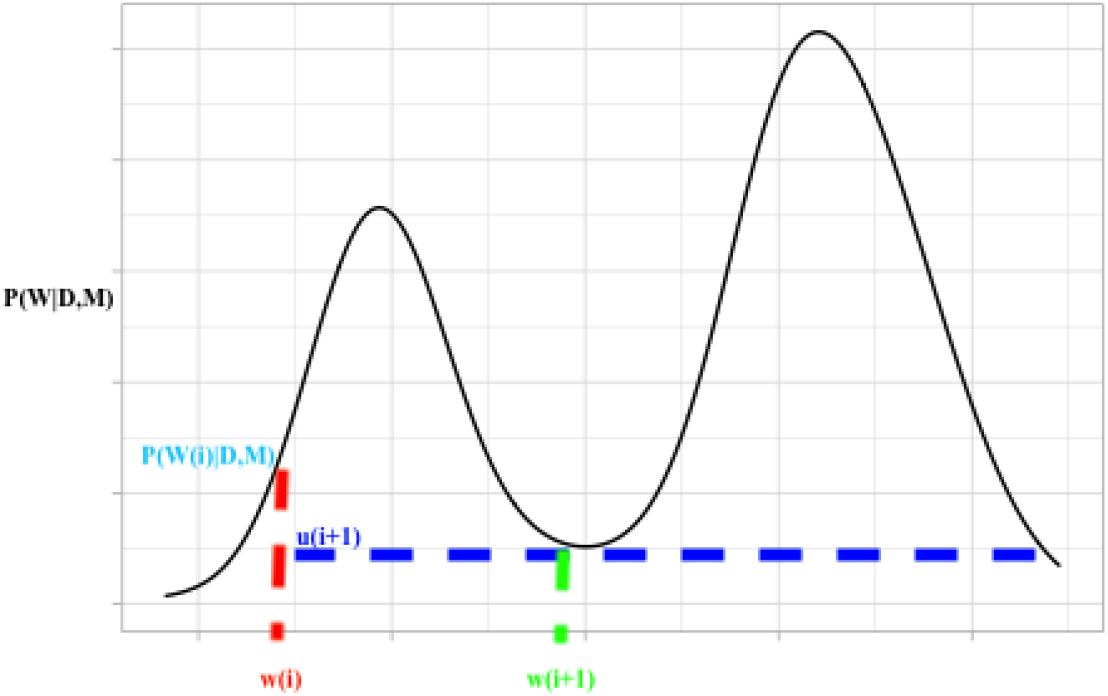
**An illustration of slice sampling, moving from a parameter sample w(i) to w(i+1) via auxiliary variable sample u(i+1)**

While sample acceptance is guaranteed with slice sampling, a large slice window can lead to computationally inefficient sampling while a small window can lead to poor mixing.

#### Hybrid Monte Carlo (HMC) and the No-U-Turn Sampler (NUTS)

Metropolis-Hastings (MH) and slice sampling suffer from excessive random walk behaviour - where the next state of the Markov Chain is randomly proposed from a proposal distribution [12, 13]. This results in inefficient sampling with low acceptance rates and small effective sample sizes.

[14] proposed HMC, which suppresses random walk behaviour by augmenting the parameter space with auxiliary momentum variables. HMC uses the gradient information of the posterior distribution to create a vector field around the current state, giving it a trajectory towards a high probability next state. The dynamical system formed by the model parameters *W* and the auxiliary momentum variables *p* is represented by the Hamiltonian *H*(*W, p*) written as follows [14]:

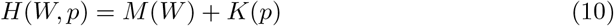

Where *M* (*W*) is the negative log-likelihood of the posterior distribution in equation 7, also referred to as the potential energy. *K*(*p*) is the kinetic energy defined by the kernel of a Gaussian with a covariance matrix *M* [15]:

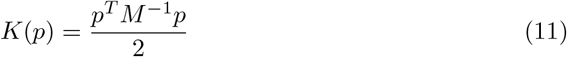

The trajectory vector field is defined by considering the parameter space as a physical system that follows Hamiltonian Dynamics. The dynamical equations governing the trajectory of the chain are then defined by Hamiltonian equations at a fictitious time *t* as follows [14]:

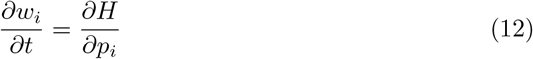

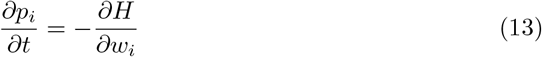

In practical terms, the dynamical trajectory is discretised using the leapfrog integrator. In the leapfrog integrator to reach the next point in the path, we take half a step in the momentum direction, followed by a full step in the direction of the model parameters - then ending with another half step in the momentum direction.

Finally due to the discretising errors that arise from the leapfrog integrator a Metropolis acceptance step is performed in order to accept or reject the new sample proposed by the trajectory[16]. In the Metropolis step the parameters proposed by the HMC trajectory *w∗* are accepted with the probability [14]:

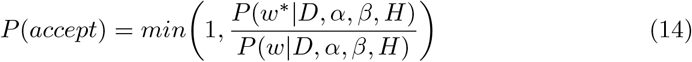

Algorithm 1 shows the pseudo-code for the HMC where *ϵ* is a discretisation stepsize.

The leapfrog steps are repeated until the maximum trajectory length *L* is reached.

##### Algorithm 1: Hybrid Monte Carlo Algorithm

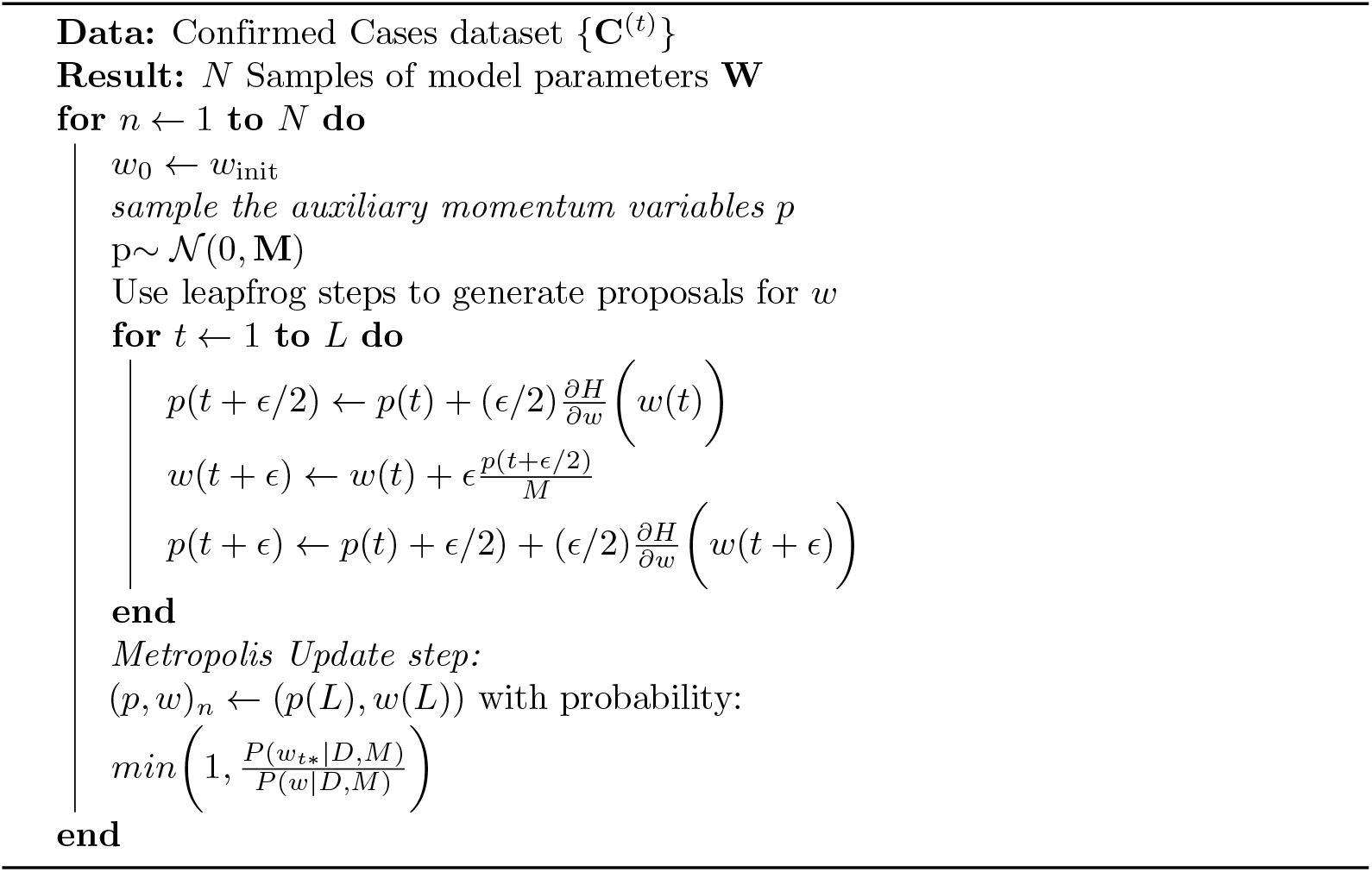

The HMC algorithm has multiple parameters that require tuning for efficient sampling, such as the step size and the trajectory length. In terms of trajectory length, a trajectory length that is too short leads to random walk behaviour similar to MH. While a trajectory length that is too long results in a trajectory that inefficiently traces back.

The stepsize is also a critical parameter for sampling, small stepsizes are computationally inefficient leading to correlated samples and poor mixing while large stepsizes compound discretisation errors leading to low acceptance rates. Tuning these parameters requires multiple time consuming trial runs.

NUTS automates the tuning of the leapfrog stepsize and trajectory length. In NUTS the stepsize is tuned during an initial burn-in phase by targeting particular levels of sample acceptance. The trajectory length is tuned by iteratively adding steps until either the chain starts to trace back (U-turn) or the Hamiltonian explodes (becomes infinite).

## Results

SIR and SEIR model parameter inference was performed using confirmed cases data upto and including 20 April 2020 and MCMC samplers described in the methodology section. Each of the samplers are run such that 2000 samples are drawn with 500 burn-in and tuning steps. We use leave-one-out(LOO) cross-validation error of [17] to evaluate the goodness of fit of each model and inference method pair.

Table 2 shows the LOO validation errors of the various models. It can be seen that the SIR model with two change points inferred by Slice Sampling displays the best model fit with the lowest mean LOO of 446.77. The SEIR model with two change points inferred using the NUTS showed a mean LOO of 454.06. We note that[4] similarly finds that the SIR model displayed superior goodness of fit to the SEIR on German data.

**Table 2.** **Leave-one out (LOO) Statistics comparing SEIR and SIR models with different number of change points using various samplers**.

| Sampler | Model | Change Points | Rank | LOO | Effective Parameters |
| --- | --- | --- | --- | --- | --- |
| SLICE | SIR | 2 | 0 | 446.77 | 9.81 |
| NUTS | SIR | 2 | 1 | 447.04 | 9.916 |
| NUTS | SEIR | 1 | 2 | 452.97 | 11.99 |
| NUTS | SEIR | 2 | 3 | 454.06 | 12.02 |
| NUTS | SEIR | 0 | 4 | 455.16 | 16.06 |
| NUTS | SIR | 1 | 5 | 462.37 | 7.66 |
| SLICE | SIR | 1 | 6 | 462.57 | 7.74 |
| MH | SIR | 2 | 7 | 475.78 | 20.52 |
| MH | SEIR | 2 | 8 | 476.41 | 10.93 |
| MH | SEIR | 1 | 9 | 494.17 | 11.41 |

We now further present detailed results of the best performing SIR Slice sampling model and SEIR with NUTS, the traceplots from these models indicating stationarity in the sampling chains are provided in appendix figures 11 and 12.

### Posterior Parameter Distributions

Figure 4 shows the posterior distributions of the SIR model parameters. The parameter estimates are *λ*_0_ ≈ 0.479 (CI[0.416, 0.554]), *λ*_1_ ≈ 0.094 (CI[0.016, 0.127]), *λ*_2_ ≈ 0.177 (CI[0.13, 0.231]), *µ* ≈ 0.136 (CI[0.096, 0.181]) and reporting delay (*D*) ≈ 6.254 (CI[4.622, 8.07]). This corresponds to *R*_0_ values of 3.522 (CI[3.06, 4.07]), 0.691 (CI[0.45, 0.93]) and 1.301 (CI[0.96, 1.70]) at the respective change points.

**Fig 4.**
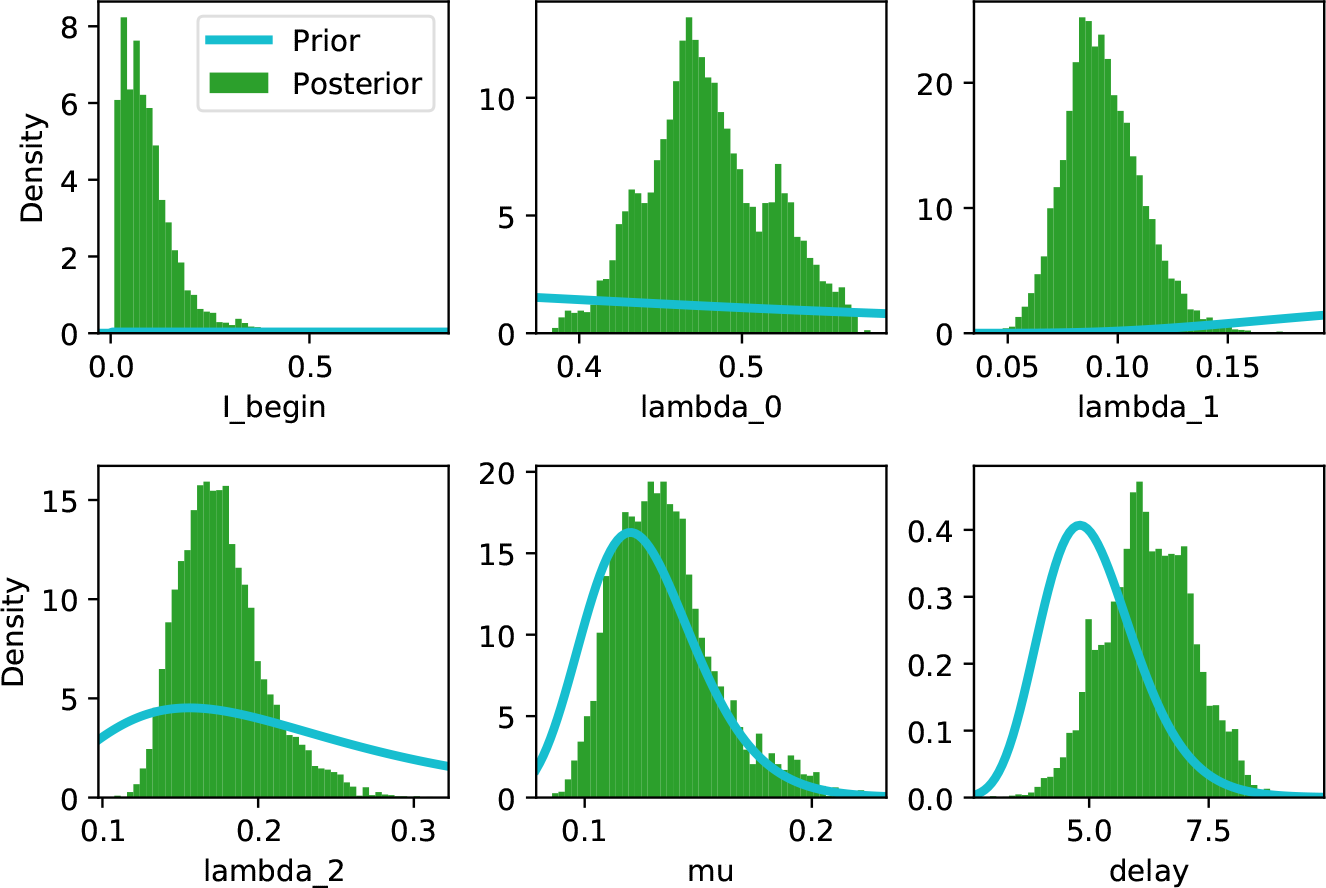
Posterior Parameter distributions for the SIR model with two change points.

Time-varying spread rates allow for inference of the impact of various state and societal interventions on the spreading rate. Figure 5 shows the fit and projections based on SIR models with zero, one and two change points. As can be seen from the plot the two change point model best captures the trajectory in the development of new cases relative to the zero and one change point models. The superior goodness of fit of the two change point model is also illustrated in table 2. The fit and projections showing similar behaviour on the SEIR model with various change points are shown in figure 6.

**Fig 5.**
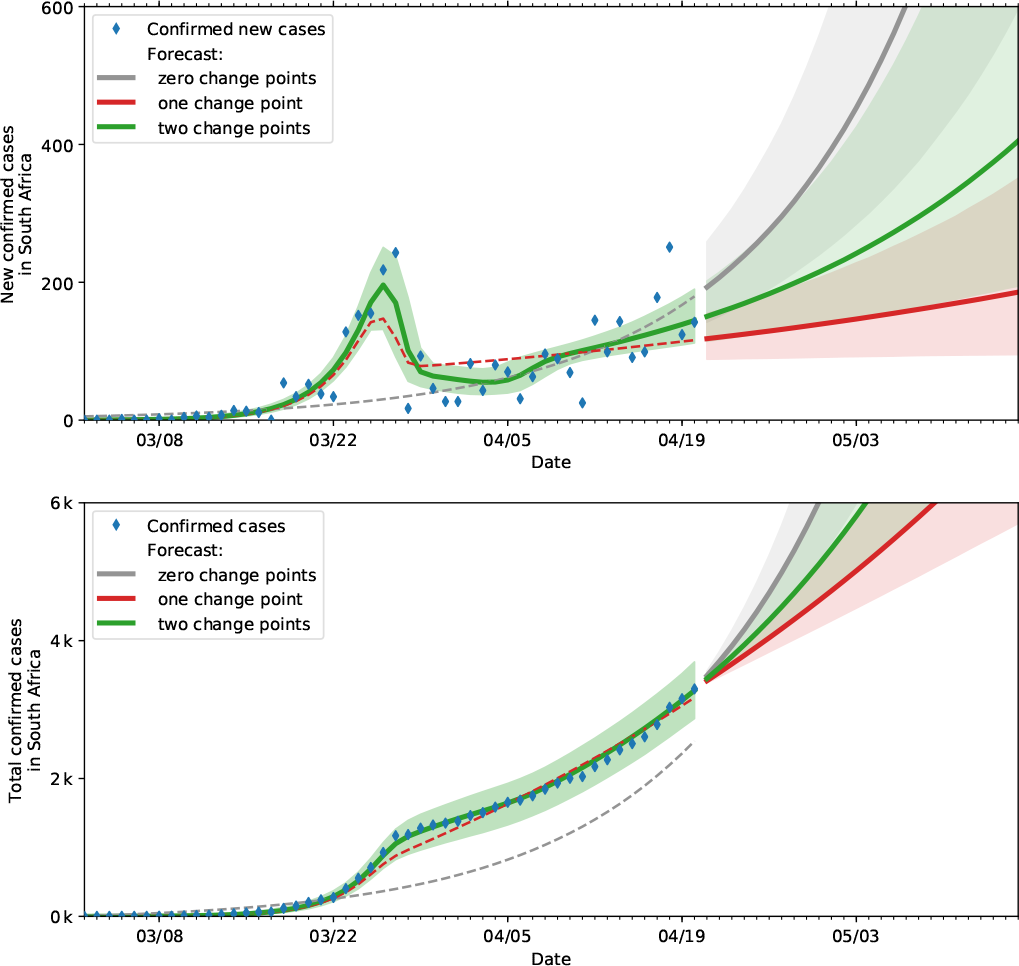
Predictions and actual data(until 20 April 2020) based on SIR models with various change points. The top plot indicates the actual and projected new cases while the bottom plot shows the actual and projected cumulative cases.

**Fig 6.**
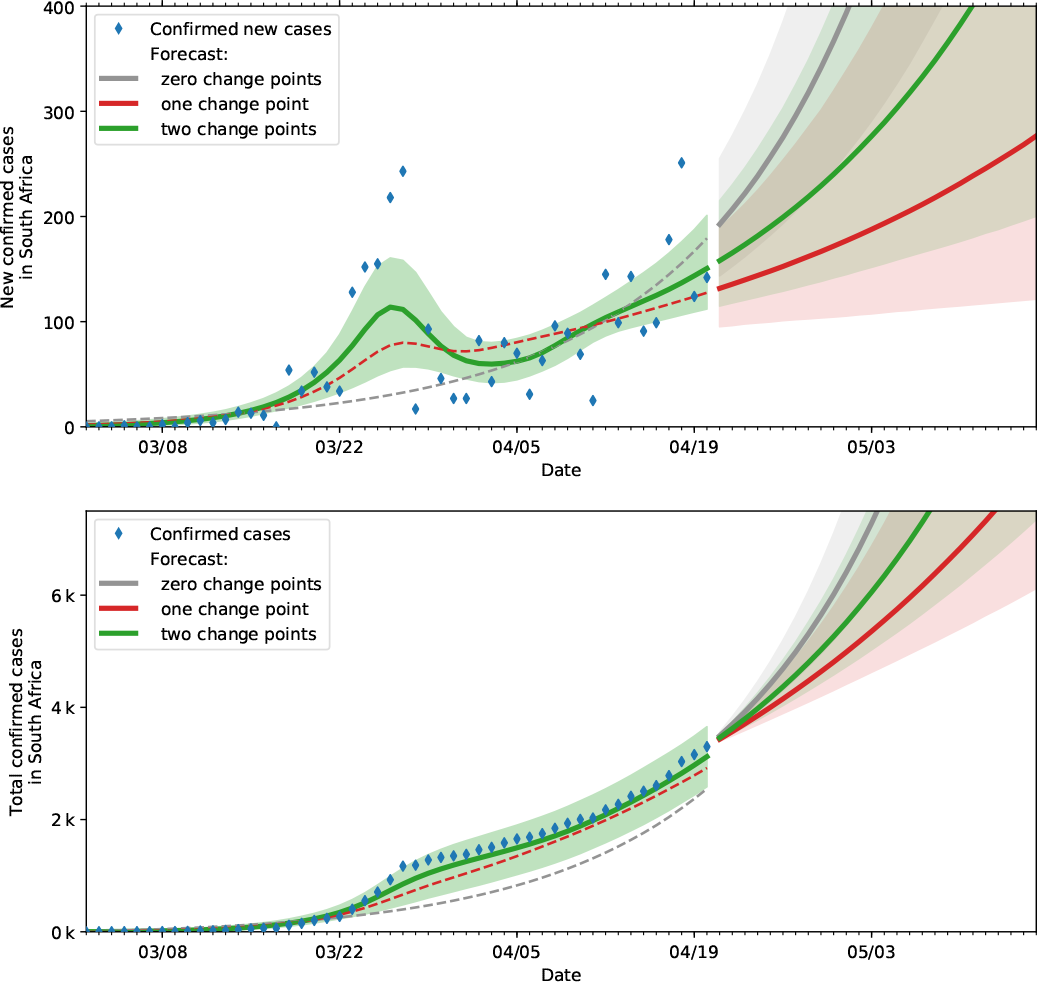
Predictions and actual data(until 20 April 2020) based on SEIR models with various change points. The top plot indicates the actual and projected new cases while the bottom plot shows the actual and projected cumulative cases.

### Reporting Delays, Incubation and Infectious period

The mean reporting delay time in days was found to be 6.254 (CI[4.622, 8.07]), literature suggests this delay includes both the incubation period and the test reporting lags. The posterior distribution incubation period from the SEIR model in figure 7 yields a median incubation period of 3.446 days (CI[1.401, 5.63]). Thus suggesting a mean laboratory reporting delay of approximately 2.8 days. A mean recovery rate *µ* ≈ 0.136 implies mean infectious period of 7.36 days which is in line with related literature [1, 4, 9].

**Fig 7.**
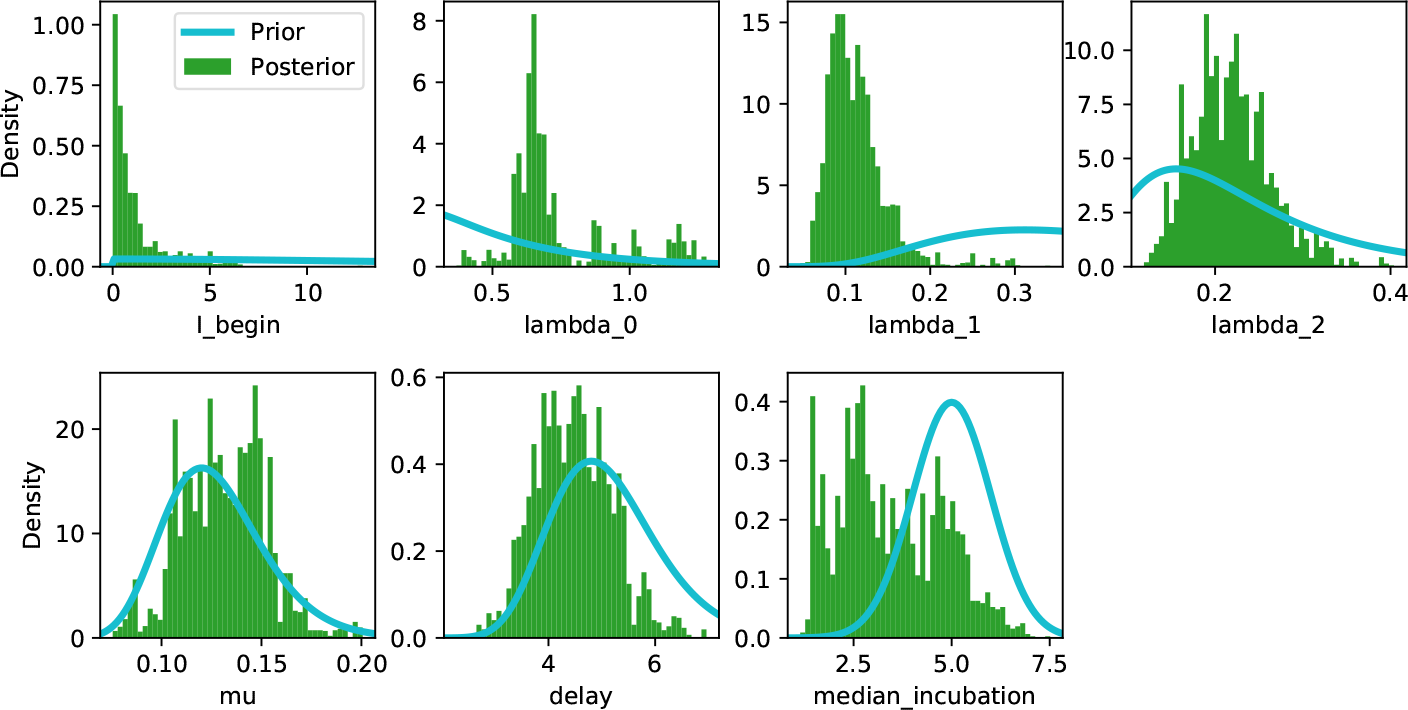
**Posterior Parameter distributions under SEIR model with two change points**.

### Timing and impact of interventions

Figure 8 depicts the posterior distributions of the spreading rates and times corresponding to each change point. We observe that the first change point is on a mean date of 18 March 2020 (CI:[16/03/2020, 20/03/2020]). This date is consistent with the travel ban, school closures and social distancing recommendations. This change point resulted in a substantial decrease in the spreading rate (80%) primarily due to the reduction in imported infections.

**Fig 8.**
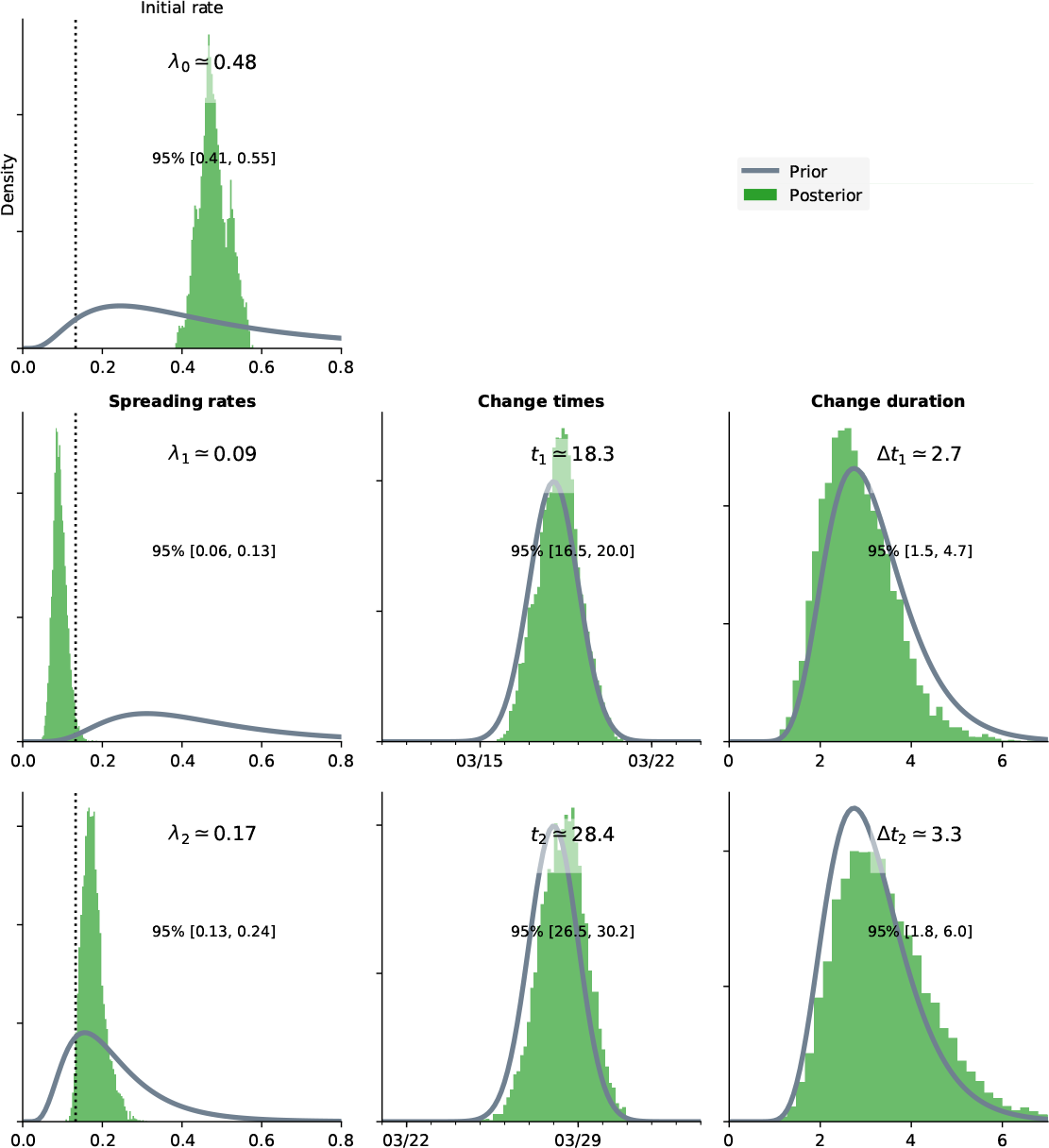
**Posterior distributions of the spreading rates(*λ*_*t*_) and the corresponding distributions of the time points.**

The second change point is observed on 28 March 2020 (CI:[26/03/2020, 30/03/2020]). This time point coincides with the announcement of mass screening and testing by the government on 30 March 2020. The resulting mean *R*_0_ of 1.30 implies a 60% decrease from the initial value.

The inference of parameters is dependent on the underlying testing processes that generate the confirmed case data. The effect of the mass screening and testing campaign was to change the underlying confirmed case data generating process by widening the criteria of those eligible for testing. While initial testing focused on individuals that either had exposure to known cases or travelled to known COVID-19 affected countries, mass screening and testing further introduced detection of community level transmissions which may contain undocumented contact and exposure to COVID-19 positive individuals.

## Discussion

We have performed Bayesian parameter inference of the SIR and SEIR models using MCMC and publicly available data as at 20 April 2020. The resulting parameter estimates fall in-line with the existing literature in-terms of mean baseline *R*_0_ (before government action), mean incubation time and mean infectious period[1, 3, 4, 9].

We find that initial government action that mainly included a travel ban, school closures and stay-home orders resulted in a mean decline of 80% in the spreading rate. Further government action through mass screening and testing campaigns resulted in a second trajectory change point. This latter change point is mainly driven by the widening of the population eligible for testing, from travellers (and their known contacts) to include the generalised community who would have probably not afforded private lab testing which dominated the initial data. This resulted in an increase of *R*_0_ to 1.301. The effect of mass screening and testing can also be seen in figure 9 indicating a mean increase in daily tests preformed from 1639 to 4374.

**Fig 9.**
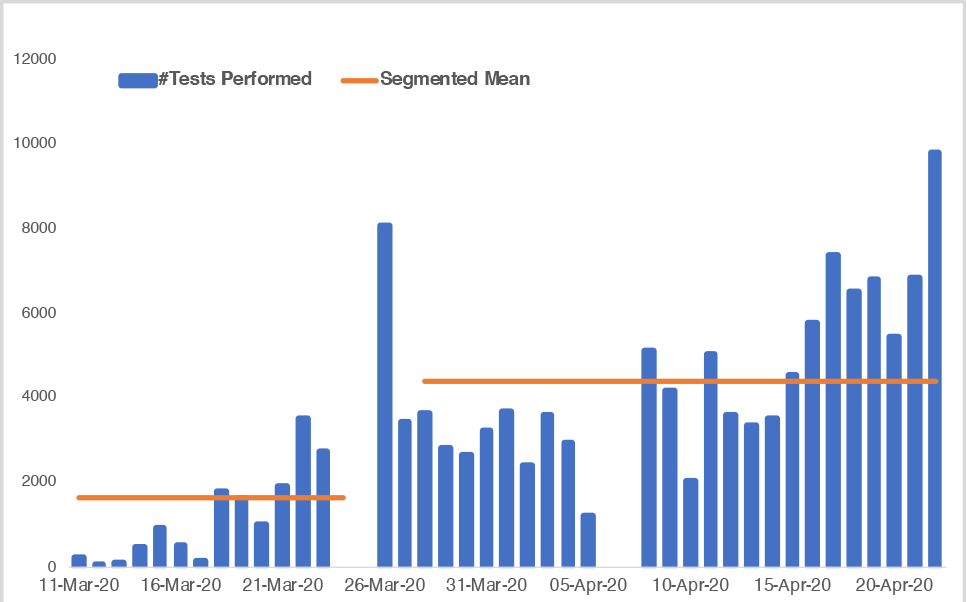
Daily COVID-19 tests performed in South Africa. The orange line indicates the segmented mean number of tests per day before and after the 28 March 2020 change point.

The second change point illustrates the possible existence of “multiple pandemics”, as suggested by [18]. Thus testing after 28 March is more indicative of community-level transmissions that were possibly not as well documented in-terms of contact tracing and isolation relative to the initial imported infection driven pandemic. This is also supported by the documented increase in public laboratory testing (relative to private) past this change point, suggesting health care access might also play a role in the detection of community-level infections[19].

## Conclusion

We have utilised a Bayesian inference framework to infer time-varying spreading rates of COVID-19 in South Africa. The time-varying spreading rates allow us to estimate the effects of government actions on the dynamics of the pandemic.

The results indicate a decrease in the mean spreading rate of 60%, which mainly coincides with the containment of imported infections, school closures and stay at home orders.

The results also indicate the emergence of community-level infections which are increasingly being highlighted by the mass screening and testing campaign. The development of the community level transmissions (*R*_0_ ≈ 1.301 (CI[0.96, 1.70])) of the pandemic at the time of publication appears to be slower than that of the initial traveller based pandemic (*R*_0_ ≈ 3.5221 (CI[3.06, 4.07])).

A future improvement to this work could include extensions to regional and provincial studies as current data suggests varied spreading rates both regionally and provincially. As more government interventions come to play priors on more change points might also be necessary.

## Data Availability

Data is publicly available

## Supporting information

**S1 Appendix**. Figures 10, 11 and 12 provide supplementary results and diagnostics from the posterior distributions.

**Fig 10.**
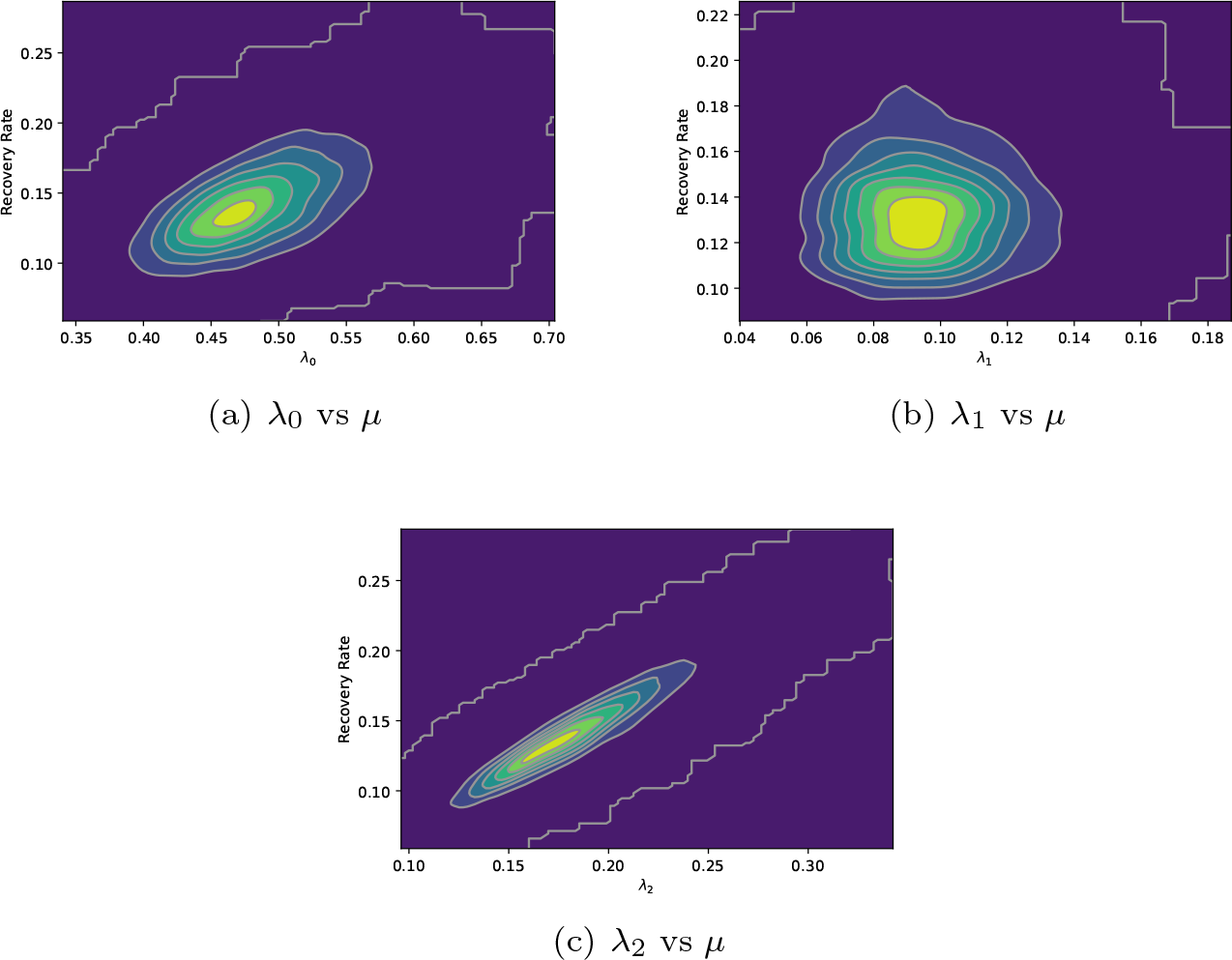
Two dimensional heat maps of the posterior distributions of the spreading rate(*λ*) and the recovery rate(*µ*) at various change points of the SIR model. The high joint density areas (in yellow) indicate likely values of *R*_0_. The baseline mean *R*_0_ estimate in 10(a) is 3.522, the first change point estimate in figure 10(b) is 0.691 while the second change point in figure 10(c) has resulted in a mean *R*_0_ estimate of 1.301.

**Fig 11.**
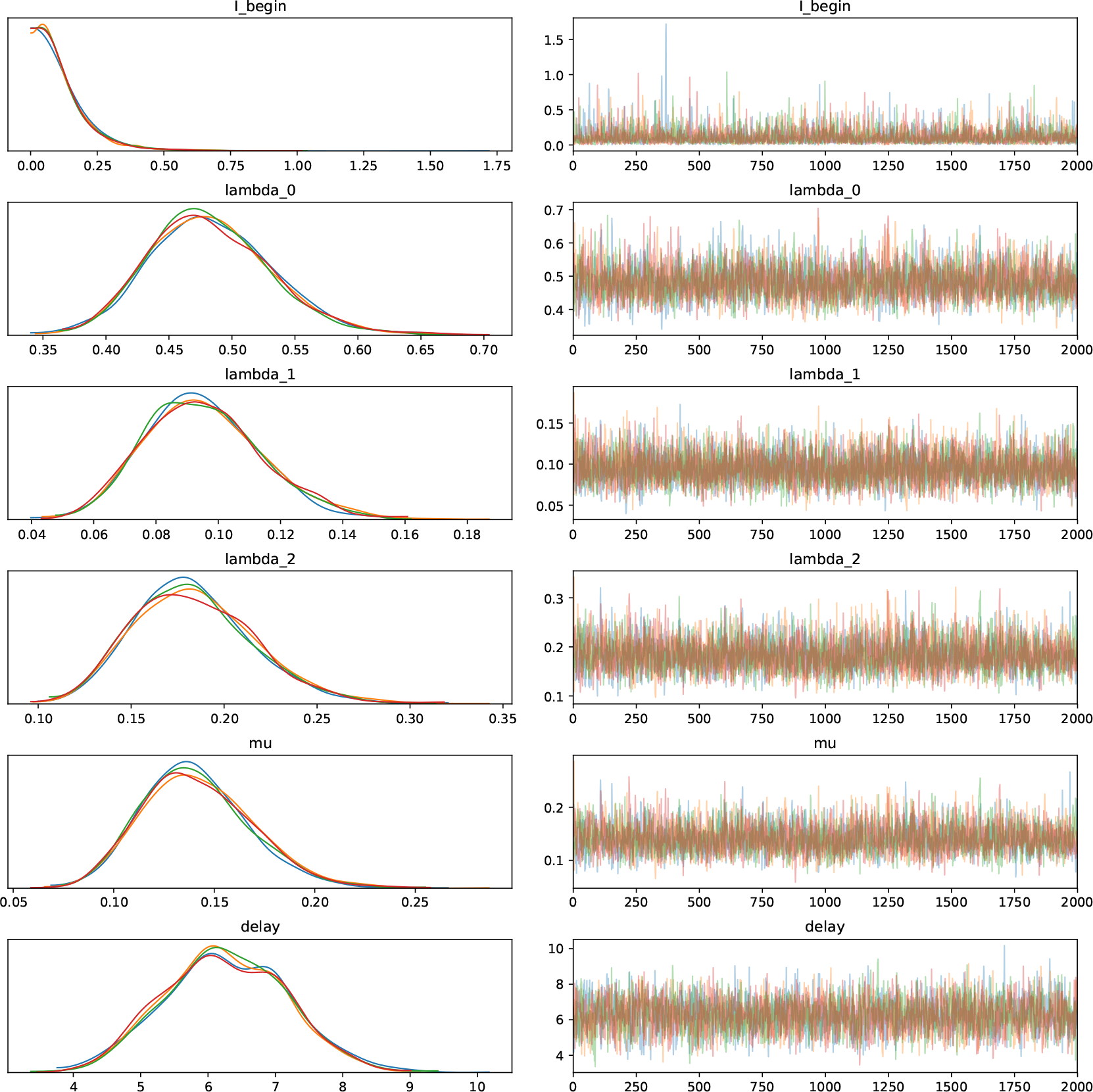
Trace plots from four concurrent chains of of the SIR model parameters inferred by Slice sampling.

**Fig 12.**
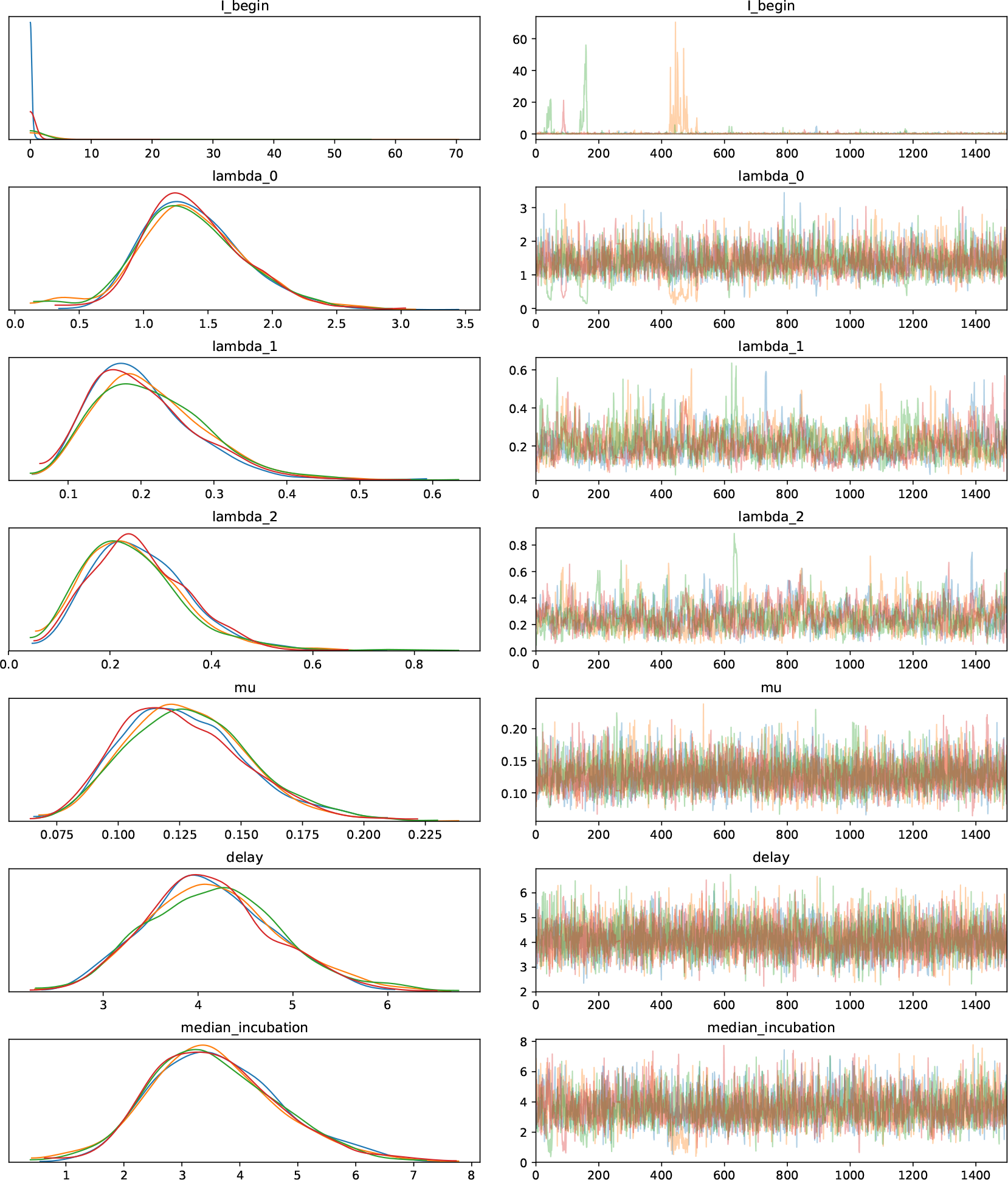
Trace plots from four concurrent chains of of the SEIR model parameters inferred by NUTS.

